# PENet: Continuous-Valued Pulmonary Edema Severity Prediction On Chest X-ray Using Siamese Convolutional Networks

**DOI:** 10.1101/2022.02.09.22270763

**Authors:** Md Navid Akbar, Xin Wang, Deniz Erdoğmuş, Sandeep Dalal

## Abstract

For physicians to take rapid clinical decisions for patients with congestive heart failure, the assessment of pulmonary edema severity in chest radiographs is vital. While deep learning has been promising in detecting the presence or absence, or even discrete grades of severity, of such edema, prediction of the continuous-valued severity yet remains a challenge. Here, we propose PENet, a deep learning framework to assess the continuous spectrum of pulmonary edema severity from chest X-rays. We present different modes of implementing this network, and demonstrate that our best model outperforms that of earlier work (mean area under the curve of 0.91 over 0.87, for nine comparisons), while saving training data and computation.

## I. Introduction

Using medical images to assess disease severity and evaluate longitudinal changes is a routine and important task in clinical decision making. For example, in the case of COVID-19 pneumonia, chest X-ray (CXR) scoring systems are used to escalate or de-escalate care, monitor treatment efficacy, and predict subsequent intubation or death [1]. In pulmonary edema, clinical decisions for patients with acute congestive heart failure (CHF) are often based on the grades of pulmonary edema severity, rather than its mere absence or presence [2]. Reliable estimation of pulmonary edema severity is challenging, since it depends on subtle findings and inter-rater agreement among even experienced radiologists is low [3].

Given the success of deep learning in computer vision, deep neural networks (DNNs) are now regularly utilized in a diverse range of medical imaging applications [4], [5]. Such DNN models have also been applied in CXRs to detect the presence of edema [6], or its discrete grades of severity [7]. These discrete grades of severity do not always reflect true continuous spectrum of change, and by discretizing, we potentially lose valuable information on continuous severity assessment. Siamese convolutional networks, already well known in the field of facial and handwriting recognition [8], have been shown recently to be effective in detecting continuous pulmonary COVID-19 severity from CXRs [9]. Inspired by this approach, our work presents PENet: a Siamese convolutional neural network to estimate the continuous scale of pulmonary edema severity in patients with CHF. To summarize our contributions, we: 1) explore weakly supervised pretraining with publicly available CXR datasets and an abnormality definition to produce continuous abnormality scores relevant to pulmonary edema without a condition specific dataset; 2) subsequently train the pretrained model with a publicly available labeled CHF dataset^1^ to predict more accurate, continuous edema severity scores; 3) train a model directly on the CHF dataset, without pretraining, and demonstrate it performs similarly to the other fully trained model with pretraining. The remainder of the paper is organized as follows: Section II outlines our model development and training techniques, Section III presents our findings and observations, and Section IV summarizes our discussion.

## II. Methods

### A. Data Collection

In this work, severity labels corresponding to the stages of edema from 4,839 individual frontal (either AP or PA) CXR images are extracted from their radiology reports, following [7]. Each CXR corresponds to an individual CHF patient from MIMIC-CXR [10], and is identified in the radiology reports under four severity levels: no edema, vascular congestion (mild edema), interstitial (moderate) edema, and alveolar (severe) edema [7]. These 4,839 labeled images are then split into train (3,354), validation (517), and test (968) splits.

### B. Preprocessing Techniques

As seen in Fig. 1, two different image preparation techniques are adopted, before feeding the input CXRs to PENet. In the first, the input CXRs are resized to 336 pixels in the shorter side and then center cropped to 320×320 pixels. In the second, the input CXRs are resized to 512 pixels in the longer side, and then symmetrically zero padded on both ends of the short side. During training, the prepared images are augmented by random translation (± 5 percent of height and width) and rotation (± 5 degrees). Finally, during all stages of training, validation, and testing, the prepared images are also mean normalized.

**Fig. 1:**
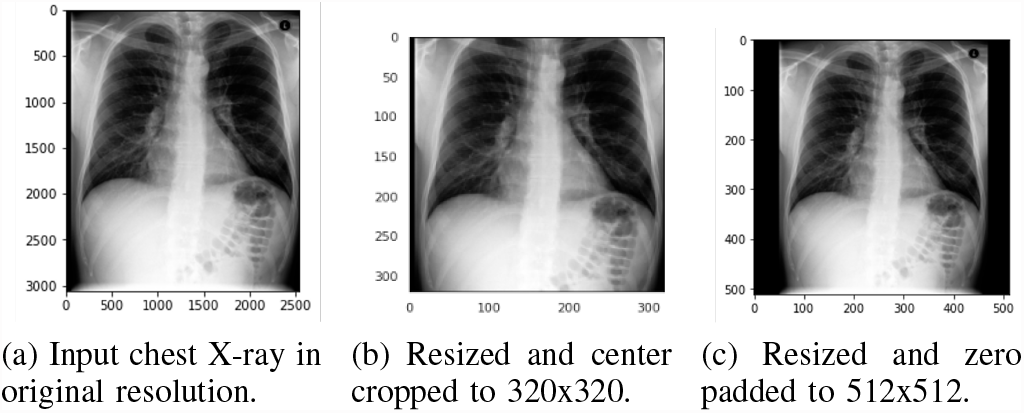
A sample patient radiograph from MIMIC-CXR [10] and two separate image preparation techniques investigated.

### C. Model Development

A convolutional Siamese neural network is used to assess the severity score of an input CXR *X*_1_, given an anchor (no edema) image *X*_2_. Both images are passed through identical parallel sub-networks *f*_*e*_() with shared weights.

The backbone of *f*_*e*_() is a DenseNet121 [11], without the last softmax layer, pretrained on ImageNet. The output of each sub-network *f*_*e*_() is a 1000-element long unnormalized vector, corresponding to the number of classes in ImageNet. Each of these vectors are then separately connected to a fully connected layer (FCL). A 9-element output vector from each FCL is then passed through a sigmoid layer to constrain each vector element in the min-max interval of [0,1]. Standard mathematical operations of elemental subtraction, square, summation, and square root are subsequently performed to obtain the Euclidean distance *d*_*e*_ between *f*_*e*_(*X*_1_) and *f*_*e*_(*X*_2_). This scoring process of a given CXR is repeated for each anchor in a pool of *k* images, and the median of the scores is recorded as either the predicted abnormality, or the edema severity score, depending on the training strategy. Fig. 2 illustrates the entire process.

**Fig. 2:**
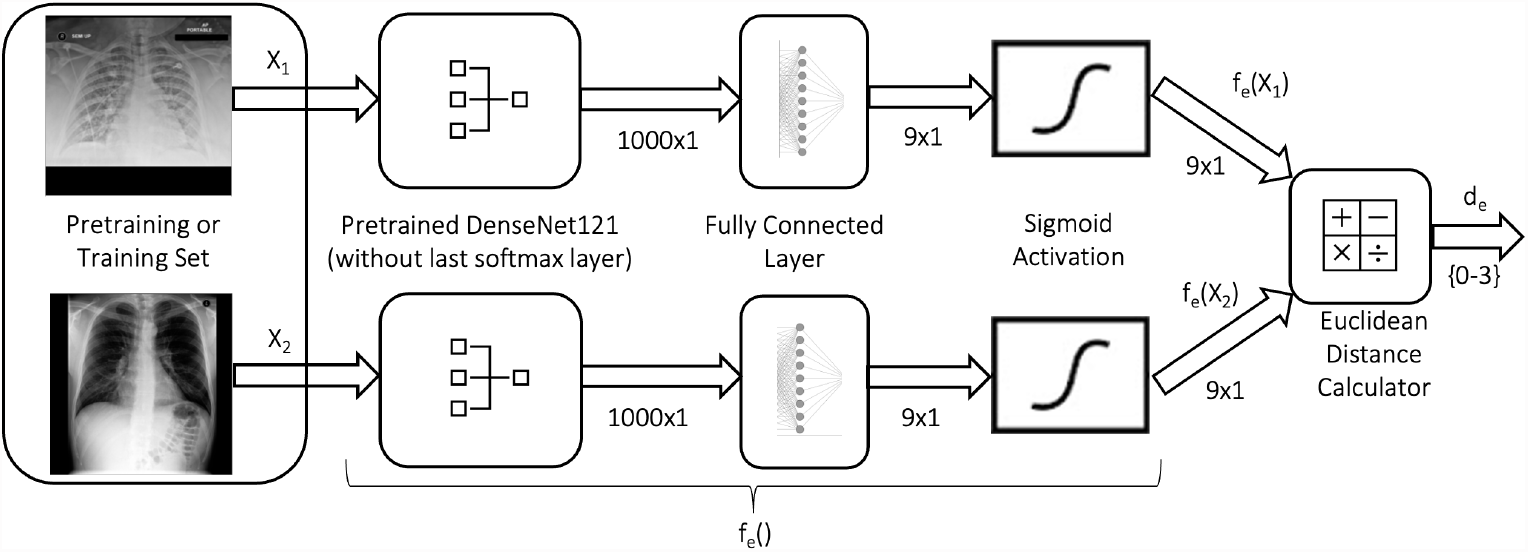
Block diagram of PENet. The scoring process is repeated for *X*_2_ ∈ {*k*} pool of anchor (no edema) images, and the median is recorded as the abnormality or the edema severity score, whichever is applicable.

### D. Abnormality Definition

For pretraining PENet further with domain CXR images, we make use of two large databases: CheXpert [12] and MIMIC-CXR [10]. Using the CheXpert labeler [12] on each radiology report associated with any image from either CheXpert or MIMIC-CXR, annotations of ’positive’, ’negative’, and ’uncertain’ are generated for several pulmonary findings [9]. To create our abnormality definition with regards to the presence of pulmonary edema or not, two board-certified radiologists were consulted. At least one positive in any of the following conditions represents an abnormality: ’lung opacity’, ’lung lesion’, ’consolidation’, ’pneumonia’, ’atelectasis’, and ’edema.’ If a report is negative for all the above conditions, or contains a ’no finding’ as an annotation summary, that image is labeled normal. All other images are treated as uncertain, and discarded from our analysis.

### E. Training Strategies

Three separate training strategies are adopted in this work. In the first, PENet is pretrained by weak supervision with CXR images from either CheXpert or MIMIC-CXR. Following the definition of abnormality outlined earlier, the images are first classified as either normal or abnormal. Image pairs used to train are then chosen as either both normal or both abnormal, or either of the two permutations of one normal and one abnormal, with an equal prior probability of selecting any of the four choices. For pretraining with MIMIC-CXR, care is also taken to discard CXRs of any subject that is common to the CHF dataset. Once this preliminary training completes, PENet is then subsequently trained on the actual smaller CHF training set. In the second strategy, regular PENet, the model is directly trained on CHF data, without pretraining on a larger CXR dataset first. In the third strategy (a variant of the second), equiprobable PENet, an equal prior probability for selecting an edema label from each severity, in both training and validation, is ensured by undersampling the overrepresented labels and oversampling the underrepresented labels.

### F. Loss and Evaluation Functions

Given input images *X*_1_ and *X*_2_, PENet calculates the Euclidean distance *d*_*e*_ between the two subnetwork outputs *f*_*e*_(*X*_1_) and *f*_*e*_(*X*_2_) as

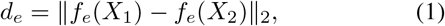

where ‖.‖_2_ denotes the Euclidean norm.

To train PENet, four loss functions are investigated. First is the contrastive loss, which minimizes *d*_*e*_ between similar images, while maximizing the distance between dissimilar images. It is given by

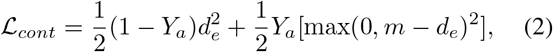

where *Y*_*a*_ = 0 if the two images are similar (both normal or abnormal) and *Y*_*a*_ = 1 if they are dissimilar (one normal and one abnormal), and *m* is the margin of dissimilarity. The weakly supervised pretraining stage uses only the contrastive loss, and *m* = 3 is chosen since it is the largest possible difference between any two severity levels.

Second is the mean square error (MSE) loss, which is given by

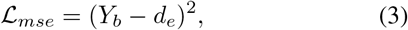

where *Y*_*b*_ ∈ {0, 1, 2, 3} indicates the severity labels.

Third, a Huber loss is also explored in training, since this loss is more robust to outliers

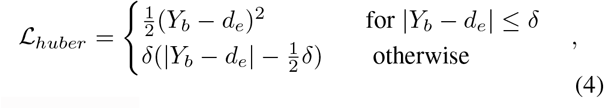

where *δ* = 1.

Finally, a combination of contrastive and MSE loss is also investigated

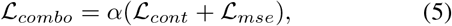

where *α*=0.5 is chosen.

For the evaluation of the trained models, the Pearson correlation coefficient *r* is calculated as

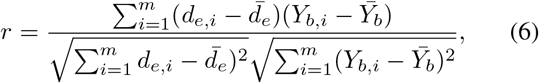

where *m* is the size of the test set, 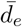 and 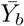 are the mean scores of the continuous-valued severity predictions and the discrete ground truth labels, respectively. Additionally, by binning the continuous *d*_*e,i*_ scores into binary comparison classes by thresholding, receiver operating characteristic (ROC) plots are generated and the areas under the ROC curves (AUCs) are recorded as indicators of performance.

### G. Experimental Details

For all training and evaluation, seed values were set to 0. While pretraining, 12,800 image pairs were chosen in training, whereas 400 pairs were chosen in validation, randomly for each epoch. During subsequent or direct training on the CHF dataset, 7,200 image pairs were chosen in training, and 800 pairs were chosen in validation, randomly for each epoch. Moreover during training and validation, the inputs are processed in mini-batches of size 8. Adam is the chosen optimizer for all models, with a learning rate of 2e-5. Model weights are saved every epoch, as long as the validation loss reduces. If the validation loss plateaus or does not improve for more than 10 epochs, early stopping is enacted.

For software, Python 3.8 was used, with support from libraries such as pytorch, scikit-learn, pandas, pickle, seaborn, matplotlib, etc. For hardware, a Ubuntu server fitted with an Intel processor and three Nvidia Maxwell GPUs is utilized for all experimentation purposes.

## III. Results and Discussions

### A. Preprocessing and Anchor Selection

In preliminary inference, the pretrained PENet models performed better with the 512×512 sized zero padded pre-processing compared to that with the 320×320 sized center cropped variant, as seen in Table I. Hence, the 512×512 preprocessing is used in all subsequent analyses. Similarly, *k*=16 anchor images from the respective validation sets of all models proved to be a good balance between performance and complexity, and is likewise chosen for all subsequent analyses.

**TABLE I:** Preliminary performance comparison between the preprocessing techniques using the best pretrained PENet models, in terms of Pearson correlation coefficient.

| Preprocessing | PENet (MIMIC-CXR) | PENet (CheXpert) |
| --- | --- | --- |
| 320x320, center crop | 0.64 ($\mathcal{L}_{combo}$) | 0.62 ($\mathcal{L}_{combo}$) |
| 512x512, zero pad | **0.72** ($\mathcal{L}_{huber}$) | **0.71** ($\mathcal{L}_{mse}$) |

Note: The loss functions used to train the individual models are indicated inside parentheses.

### B. Abnormality vs. Edema Scoring

Fig. 3 illustrates both the abnormality and edema scores for a sample patient CXR. While the fully trained PENet predicts a more accurate edema score, the weakly supervised model pretrained with MIMIC-CXR is able to generate a reasonably close abnormality score: a desirable feature for tasks such as radiology workflow prioritization [14].

**Fig. 3:**
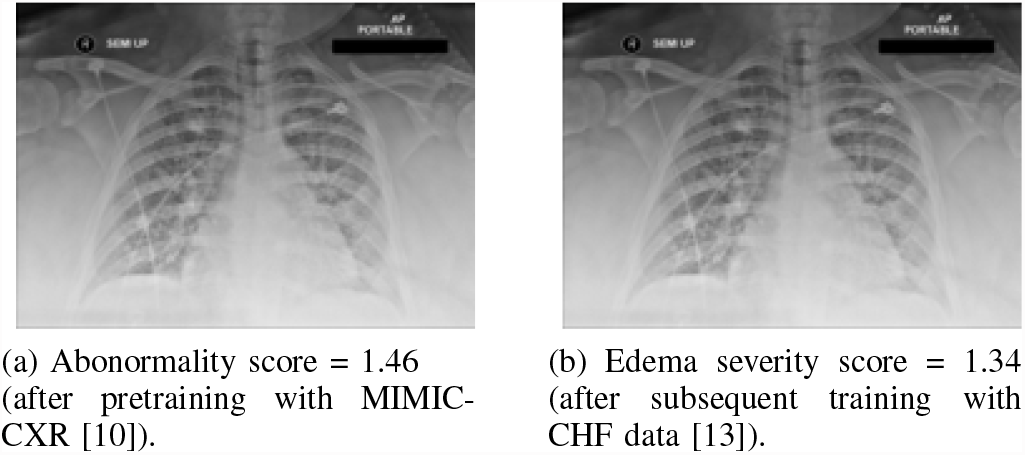
Output scores from PENet for the two-step training strategy, on a sample patient chest X-ray with ground truth severity of level 1. The fully trained model produces a more accurate prediction, but the weakly supervised pretrained model also estimates a reasonable score.

### C. Performance Across Models

Boxplots in Fig. 4 outline the best individual performances of the four PENet variants: the two models additionally pre-trained on large CXR datasets (MIMIC-CXR and CheXpert), and the two models with direct CHF dataset training. All the plots show a common linear trend of the continuous-valued predicted edema severity with the discrete ground truth edema severity. Interestingly, it is observed that the PENet pretrained on MIMIC-CXR and CheXpert datasets exhibit a similar performance as does the regular PENet without any CXR pretraining, in terms of correlation. This indicates that the computationally expensive CXR pretraining might not have provided any noticeable benefit in our experimental scenario. This phenomenon may have two possible explanations. First, pretrained on CXRs or not, all models use DenseNet121 as the network backbone, which had been pretrained on ImageNet. Second, PENet works on pairs of images. Likewise, even when the actual training set is *n*, PENet can take

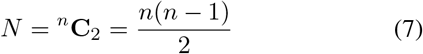

possible combinations of pairs as inputs, where *N* is the size of the synthetic training set. Thus, the maximum size of *N* PENet can distinguish becomes a large number: about 5.6 million in our case.

**Fig. 4:**
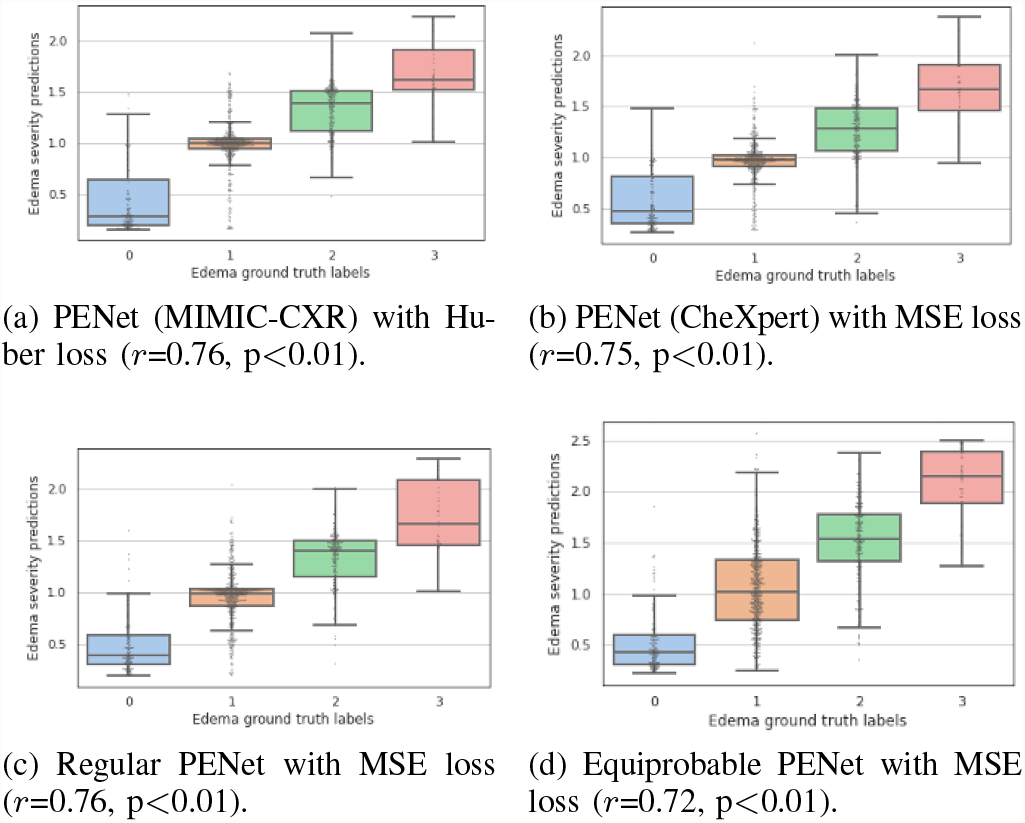
Boxplots showing the continuous-valued edema score outputs from different variants of PENet, against the discrete ground truth severity labels. The boxes indicate the median and interquartile range (IQR), with whiskers extending to points within 1.5 IQRs of the IQR boundaries. Regular PENet trained directly with CHF data [13] performs as well as PENet pretrained with MIMIC-CXR [10] or CheXpert [12]. Equiprobable PENet has a shift in the distribution of its predictions, with a slight overall drop in performance.

### D. ROC Analysis

The ROC curves of the two best PENet variants can be plotted as seen in Fig. 5, for nine separate binary comparisons. The thresholds chosen for discretizing the continuous predictions into categorical labels are [0≤ *d*_*e*_*<*0.5], [0.5≤ *d*_*e*_*<*1.5], [1.5≤ *d*_*e*_*<*2.5] and [2.5≤ *d*_*e*_ ≤] for labels 0, 1, 2, and 3, respectively. As expected, both models performed almost impeccably on the task of distinguishing images spaced farthest along the level of severity (e.g. 0 vs. 3), while they struggled the most on the tasks of classifying between adjacent states (e.g. 2 vs. 3). There is no statistical difference in performance at 5% level of significance.

**Fig. 5:**
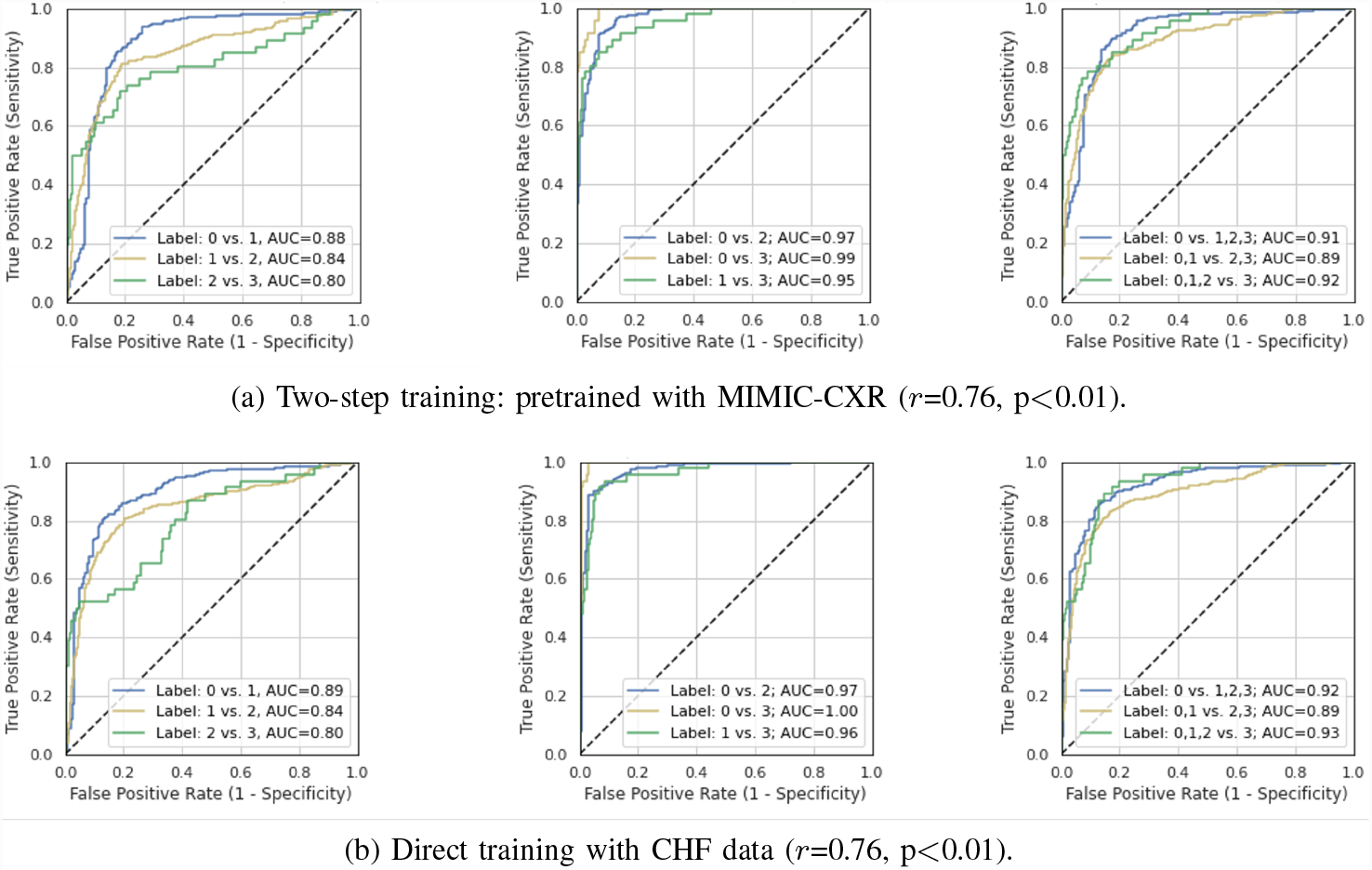
Receiver operating characteristic (ROC) plots. PENet trained directly with the CHF data [13] has similar area under ROC curve (AUC) performance, compared to when additionally pretrained with MIMIC-CXR, with no statistical difference at 5% level of significance.

### E. Comparison with Previous Work

Table II compares the performance of PENet with the results from [7]. It is observed that the regular PENet beats both the earlier ImageNet trained model, as well as the computationally heavy semisupervised model (pretrained on MIMIC-CXR, in a higher resolution), in seven out of nine AUC comparisons. The equiprobable PENet does better on the 2 vs. 3 and 0,1,2 vs. 3 comparisons, but performs slightly worse overall compared to the regular model. In the latter, the spread and accuracy of prediction for level 3 edema (fewest training samples, thus oversampled) improves, while that of level 1 (most training samples, thus undersampled) declines.

**TABLE II:** Comparison of area under the curve (AUC) performance with prior work.

| Comparison | Semisupervised* [7] | ImageNet Trained [7] | PENet (regular) | PENet (equiprobable) |
| --- | --- | --- | --- | --- |
| 0 vs. 1 | 0.79 | 0.66 | **0.89** | 0.87 |
| 0 vs. 2 | 0.88 | 0.81 | **0.97** | **0.97** |
| 0 vs. 3 | 0.99 | 0.87 | **1.00** | **1.00** |
| 1 vs. 2 | 0.69 | 0.73 | **0.84** | 0.79 |
| 1 vs. 3 | 0.93 | 0.82 | **0.96** | **0.96** |
| 2 vs. 3 | 0.88 | 0.63 | 0.80 | 0.86 |
| 0 vs. 1,2,3 | 0.85 | 0.74 | **0.92** | 0.90 |
| 0,1 vs. 2,3 | 0.88 | 0.81 | **0.89** | 0.86 |
| 0,1,2 vs. 3 | 0.96 | 0.82 | 0.93 | 0.94 |

Note: Current work uses the same training samples (3354), but more test samples (968 vs. 141).
\* Pretrained on MIMIC-CXR [10], using 2048x2048 sized input images. All other models each use DenseNet121 [11] (without classification layer, pretrained on ImageNet) as the network backbone, with 512x512 resolution input images.

## IV. Conclusion and Future Work

In this work, we presented PENet, a Siamese convolutional network to assess the severity of pulmonary edema from chest radiographs. Using an abnormality definition and a general chest X-ray dataset, our weakly supervised model is able to assess continuous-valued abnormality scores for edema, without the need for an edema specific dataset. When subsequently, or directly, trained on an edema dataset with discrete labels, PENet can predict continuous-valued severity scores with greater accuracy. For severity score prediction, we found that pretraining appears to provide no additional benefit in this particular task, thus potentially saving valuable training samples and the need for an abnormality definition. This is likely a consequence of the large number of synthetic image pairs PENet is able to extract for training from a relatively smaller dataset. Directly trained regular PENet even outperforms the best performing semisupervised model from earlier work, using less training data and lower resolution input images. As future work, we would like to perform cross-validation, and visualize the regions of interest for PENet.

## Data Availability

This study involves publicly available human data, which can be obtained from: https://physionet.org/content/mimic-cxr-pe-severity/1.0.1/

## ACKNOWLEDGMENT

We would like to thank Dr. Steven Horng and Dr. Seth J. Berkowitz (Beth Israel Deaconess Medical Center, Harvard Medical School, Boston, MA, USA) for their guidance in preparing the abnormality definition. We would also like to thank Ruizhi Liao, PhD (Department of Electrical Engineering and Computer Science, Massachusetts Institute of Technology) for providing us the edema labels.

## Footnotes

1 https://physionet.org/content/mimic-cxr-pe-severity/1.0.1/

## References

[1] J. P. Cohen, L. Dao, K. Roth, P. Morrison, Y. Bengio, A. F. Abbasi, B. Shen, H. K. Mahsa, M. Ghassemi, H. Li et al., “Predicting covid-19 pneumonia severity on chest x-ray with deep learning,” Cureus, vol. 12, no. 7, 2020.

[2] M. Gheorghiade, F. Follath, P. Ponikowski, J. H. Barsuk, J. E. Blair, J. G. Cleland, K. Dickstein, M. H. Drazner, G. C. Fonarow, T. Jaarsma et al., “Assessing and grading congestion in acute heart failure: a scientific statement from the acute heart failure committee of the heart failure association of the european society of cardiology and endorsed by the european society of intensive care medicine,” European journal of heart failure, vol. 12, no. 5, pp. 423–433, 2010.

[3] M. Hammon, P. Dankerl, H.L. Voit-Höhne, M. Sandmair, F. J. Kammerer, M. Uder, and R. Janka, “Improving diagnostic accuracy in assessing pulmonary edema on bedside chest radiographs using a standardized scoring approach,” BMC anesthesiology, vol. 14, no. 1, pp. 1–9, 2014.

[4] J. Liu, Y. Pan, M. Li, Z. Chen, L. Tang, C. Lu, and J. Wang, “Applications of deep learning to mri images: A survey,” Big Data Mining and Analytics, vol. 1, no. 1, pp. 1–18, 2018.

[5] M. N. Akbar, M. Yarossi, M. Martinez-Gost, M. A. Sommer, M. Dannhauer, S. Rampersad, D. Brooks, E. Tunik, and D. Erdog?muş, “Mapping motor cortex stimulation to muscle responses: a deep neural network modeling approach,” in Proceedings of the 13th ACM International Conference on PErvasive Technologies Related to Assistive Environments, 2020, pp. 1–6.

[6] P. Rajpurkar, J. Irvin, K. Zhu, B. Yang, H. Mehta, T. Duan, D. Ding, A. Bagul, C. Langlotz, K. Shpanskaya, M. P. Lungren, and A. Y. Ng, “Chexnet: Radiologist-level pneumonia detection on chest x-rays with deep learning,” pp. 3–9, 2017. [Online]. Available: http://arxiv.org/abs/1711.05225

[7] S. Horng, R. Liao, X. Wang, S. Dalal, P. Golland, and S. J. Berkowitz, “Deep learning to quantify pulmonary edema in chest radiographs,” Radiology: Artificial Intelligence, vol. 3, no. 2, p. e190228, 2021.

[8] D. Chicco, “Siamese neural networks: An overview,” Artificial Neural Networks, pp. 73–94, 2021.

[9] M. D. Li, N. T. Arun, M. Gidwani, K. Chang, F. Deng, B. P. Little, D. P. Mendoza, M. Lang, S. I. Lee, A. O’Shea et al., “Automated assessment and tracking of covid-19 pulmonary disease severity on chest radiographs using convolutional siamese neural networks,” Radiology: Artificial Intelligence, vol. 2, no. 4, p. e200079, 2020.

[10] A. E. Johnson, T. J. Pollard, N. R. Greenbaum, M. P. Lungren, C.-y. Deng, Y. Peng, Z. Lu, R. G. Mark, S. J. Berkowitz, and S. Horng, “Mimic-cxr-jpg, a large publicly available database of labeled chest radiographs,” arXiv preprint 1901.07042, 2019.

[11] G. Huang, Z. Liu, L. Van Der Maaten, and K. Q. Weinberger, “Densely connected convolutional networks,” in Proceedings of the IEEE conference on computer vision and pattern recognition, 2017, pp. 4700–4708.

[12] J. Irvin, P. Rajpurkar, M. Ko, Y. Yu, S. Ciurea-Ilcus, C. Chute, H. Marklund, B. Haghgoo, R. Ball, K. Shpanskaya et al., “Chexpert: A large chest radiograph dataset with uncertainty labels and expert comparison,” in Proceedings of the AAAI conference on artificial intelligence, vol. 33, no. 01, 2019, pp. 590–597.

[13] R. Liao, D. Moyer, M. Cha, K. Quigley, S. Berkowitz, S. Horng, P. Golland, and W. M. Wells, “Multimodal representation learning via maximization of local mutual information,” arXiv preprint 2103.04537, 2021.

[14] I. Baltruschat, L. Steinmeister, H. Nickisch, A. Saalbach, M. Grass, G. Adam, T. Knopp, and H. Ittrich, “Smart chest x-ray worklist prioritization using artificial intelligence: a clinical workflow simulation,” European radiology, vol. 31, no. 6, pp. 3837–3845, 2021.

